# Association of Biological Age with Tumor Microenvironment in Patients with Esophageal Adenocarcinoma

**DOI:** 10.1101/2022.11.14.22282214

**Authors:** C.J. Ravensbergen, Y. van Holstein, S.C. Hagenaars, A.S.L.P. Crobach, S. Trompet, J.E.A. Portielje, N.A. de Glas, D. van Heemst, F. van den Bos, R.A.E.M. Tollenaar, W.E. Mesker, S.P. Mooijaart, M. Slingerland

## Abstract

Biological age-related adaptations have been shown to modulate the non-malignant cells comprising the tumor microenvironment (TME). In the current work, we studied the association between biological age and TME characteristics in patients with esophageal adenocarcinoma. We comparatively assessed intratumoral histologic stroma quantity, tumor immune cell infiltrate, and blood leukocyte and thrombocyte count in 72 patients stratified in 3 strata of biological age (younger <70 years, fit older ≥70 years, and frail older adults ≥70 years), as defined by a geriatric assessment. Frailty in older adults was predictive of decreased intratumoral stroma quantity (B = -14.66% stroma, *P* = 0.022) relative to tumors in chronological-age-matched fit older adults. Moreover, in comparison to younger adults, frail older adults (*P* = 0.032), but not fit older adults (*P* = 0.302), demonstrated a lower blood thrombocyte count at the time of diagnosis. Lastly, we found an increased proportion of tumors with a histologic desert TME phenotype in frail older adults. Our findings provide a biological underpinning for the clinical relevance of assessing frailty in patients with esophageal adenocarcinoma, further justifying the need for standardized geriatric assessment in geriatric cancer patients.

## Introduction

Esophageal cancer is the seventh most common cancer worldwide and typically tends to manifest at an older age, with a median age of 68-70 years at the time of diagnosis (1, 2). Despite improvements in treatment regimens, the overall prognosis remains poor with an approximate 5-year survival rate of 20% (3, 4). Survival rates are worse in older patients compared to younger patients and this survival discrepancy has increased over the past recent years (5). Marked heterogeneity in time-dependent functional decline in older adults results in varying grades of clinically manifest patient fitness or frailty (6, 7). As opposed to chronological age, the concept of biological age aims to account for this health heterogeneity by integrating personalized clinical and molecular patient characteristics (8). Remarkably, despite 60% of the patients being over 65 years of age at the time of diagnosis, current treatment guidelines for esophageal cancer do not discriminate between fit and frail older adults (3). Both chronological age and frailty are associated with higher postoperative complication rates and mortality, and chemotherapy toxicity (9-11). In addition, we recently reported a high prevalence of geriatric deficits in older patients with curable esophageal cancer that was subsequently associated with high chemoradiotherapy discontinuation rates (12).

The tumor microenvironment (TME) comprises a dynamic ecosystem of cellular (i.e., immune cells, stroma cells) and non-cellular (i.e., extracellular matrix) components that surround neoplastic cells (13, 14). Our group and others have previously demonstrated the clinical significance of assessing histologic TME parameters, such as the tumor-stroma ratio (TSR) and tumor immune cell infiltration, in esophageal cancer (15-19). A lower TSR reflects high intratumoral stroma quantity and is associated with poor patient prognosis and pathologic response to neoadjuvant chemoradiotherapy in esophageal squamous cell and adenocarcinoma (15, 17). In contrast, increased tumor infiltration of immune cell subsets is correlated to a favorable patient prognosis in esophageal cancer (18, 19). Coinciding with local histology, blood cell counts in the circulation were recently shown to be associated with distinct histologic TME characteristics (20-23). A report on breast cancer demonstrated increased blood thrombocyte counts in patients with stroma-rich tumors (20). In addition, blood leukocyte counts were found to be correlated to tumor-infiltrating immune cells (21-23). Circulatory blood cell count profiles may therefore reflect local TME features in patients with esophageal cancer. While age-related adaptations have been shown to modulate the TME, it remains unclear whether age-related re-programming of the TME is a result of the biological aging process, or merely dependent on chronological age (24).

In the current work, we aimed to study the association between biological age and the TME in patients with esophageal adenocarcinoma. To this aim, we compared intratumoral stroma quantity, histologic tumor immune cell infiltrate, and blood leukocyte and thrombocyte count over strata of biological age (younger adults <70 years, fit older adults ≥70 years, and frail older adults ≥70 years), as defined by a geriatric assessment (GA).

## Methods

### Patients and tissue material

The patient cohort consisted of patients with esophageal adenocarcinoma, clinical stage I–IV, that underwent treatment at the Leiden University Medical Center or affiliated hospitals (Haaglanden Medisch Centrum, Alrijne Zorggroep, Groene Hart Ziekenhuis and Tergooi Medisch Centrum) between 2010 and 2019. The cohort was categorized into two main groups based on the cut-off age of 70 years. The patient population aged <70 years was part of a study, previously published by our group (17). The patient population aged ≥70 years is concurrently part of an ongoing prospective cohort study, the *Triage of Elderly Needing Treatment* (TENT) study. More details on this study were described elsewhere (25). Patients were diagnosed via esophagogastroduodenoscopy with histologic confirmation. Due to the small number of available patients, no inclusion criteria were based on tumor stage (American Joint Committee on Cancer Staging, 8^th^ edition) or treatment regimens. Clinical data, pathology reports, and blood leukocyte and thrombocyte counts at the time of diagnosis were retrospectively collected from the electronic patient files. Hematoxylin and eosin (H&E) stained pre-treatment primary tumor biopsies were collected. In the case of referred patients, the original biopsy slides were collected from regional hospitals using the Dutch national Pathology Registry (PALGA) (26). All tissue samples were coded and handled according to ethical standards (‘Code for Proper Secondary Use of Human Tissue’, Dutch Federation of Medical Scientific Societies). This study was approved by the Medical Ethics Committee of the LUMC (ID number NL53575.058.15). All participants or a proxy provided written informed consent.

### Geriatric assessment and frailty definition

Patients included in the TENT study underwent a geriatric assessment (GA) based on the four geriatric domains: the social, somatic, psychological, and functional domain. Patients with a deviant score on ≤1 geriatric domain were classified as fit. Patients who scored abnormally on at least two domains were classified as frail. A domain was considered abnormal if at least one test of the corresponding domain was scored abnormal. The social domain was assessed by asking about the current living situation and was considered abnormal when patients were institutionalized. The somatic domain contained comorbidity, polypharmacy, and malnutrition assessed by the Mini Nutritional Assessment Short Form (MNA-SF®; range 0 to 14, cutoff score ≤11) (27). Comorbidities were assessed by the Charlson Comorbidity Index (CCI) (28). The somatic domain was abnormal in the case of CCI ≥1 point (on top of points for solid tumor presence), the number of medications ≥5, and/or MNA-SF® ≤11. The psychological domain included a history of delirium, dementia, and cognitive impairment according to the Six-Item Cognitive Impairment Test (6CIT; range 0 to 28, cut-off score ≥8) (29). The functional domain was abnormal in case of a fall incident in the past six months or functional dependency according to the Katz Activities of Daily Living (ADL) questionnaire (range 0 to 6; cut-off score ≥ 2) or Lawton Instrumental Activities of Daily Living (IADL) (range 0 to 5 for men, cut-off score ≤ 4; range 0 – 8 for women, cut-off score ≤ 7) (30, 31).

### Digital Scanning of histology slides

Diagnostic H&E-stained biopsy samples were digitally scanned using the Pannoramic 250 scanner (3DHISTECH, Budapest, Hungary) at ×20 magnification (0.39 µm per pixel) and stored as .mrxs files.

### Visual scoring of the tumor-stroma ratio and tumor immune cell infiltrate

The TSR was scored on digitalized routine histology hematoxylin and eosin (H&E)-stained slides of pre-treatment biopsies in CaseViewer software (3DHISTEC, Budapest, Hungary). The tumor area with the highest amount of stroma was selected, as described previously in esophageal adenocarcinoma (15). A detailed description of the methodology and scoring eligibility criteria can be found in the protocol for standardized TSR scoring (32). In short, the percentage of stroma was scored in increments per ten percent, and the tumor was subsequently categorized as a stroma-high (>50% stroma) or stroma-low (≤50% stroma) tumor. All slides were independently scored by two observers (C.R. & S.H.). When consensus could not be reached, the assessment of a third observer (A.C., board-certified pathologist) was decisive.

The tumor immune cell infiltrate (TICI) was defined as the infiltrating immune cells within the tumor stroma and scored in accordance with the International Immuno-Oncology Biomarker Working Group guidelines for tumor-infiltrating lymphocyte (TIL) assessment in solid tumors (33, 34). Full sections of the biopsy samples were used to obtain a full assessment of the average stromal immune cell infiltration. Some adaptations to the standardized working group protocol were made. First, in line with our previous report, no distinction was made between infiltrating immune cell subsets (35). Therefore, tumor immune cell infiltrate, as defined in this study, comprised all inflammatory cells (i.e., mononuclear and polymorphonuclear cells) as opposed to mononuclear cells exclusively. In addition, given the challenging nature of immune infiltrate scoring, the percentage of immune cell infiltration was scored in increments per ten percent as opposed to a continuous scale of single-digit percentages. Tumor immune infiltrate scoring currently lacks a validated cut-off value for risk stratification. In conjunction with other publications on immune cell scoring in esophageal cancer, we applied the median immune cell infiltrate percentage of our cohort as a cut-off value, where an immune cell infiltrate greater than the median percentage was defined as immune-high and an immune cell infiltrate less than or equal to the median percentage was defined as immune-low (18, 19).

### Statistical analysis

The R programming language (version 4.0.5; https://www.r-project.org/) was used for statistical analyses and data visualization (packages tidyverse, corrplot, ggfortify, ggExtra, and viridis). Variable distribution was assessed with the Shapiro-Wilk test, followed by either parametric or non-parametric testing. The Fisher’s exact test or the Chi-squared test was used for categorical variables. For continuous variables, the Mann-Whitney test was used and the median value and interquartile range were reported. Linear regression analysis was used to test the statistical significance of categorical and continuous predictor variables on continuous outcome variables. Interobserver variability was evaluated with Cohen’s Kappa coefficient (*Κ*). A two-tailed *P*-value of <0.05 was considered statistically significant.

## Results

### Cohort Selection, Patient Characteristics, and Histopathology Features

The initial total cohort of 76 esophageal adenocarcinoma patients consisted of three groups, a younger adult (<70 years, *n* = 38) group, a fit older (≥70 years, *n* = 19) group, and a frail older group (≥70 years, *n* = 19). Out of the 76 patients in the cohort, a total of 72 H&E-stained slides proved to be of sufficient quality for histopathologic analysis and were included for further analysis (*Supplementary Figure S1*). The baseline tumor and geriatric characteristics of the 72 eligible patients can be found in *Table 1*.

**Table 1.** Baseline characteristics of patients with esophageal adenocarcinoma. The ≥70y population is further stratified into fit or frail groups.

| Geriatric Characteristics | Younger adult (<70y)<br>n = 35 | Fit older adult ($\geq 70$y)<br>n = 19 | Frail older adult ($\geq 70$y)<br>n = 18 | P-value (younger adult vs. fit older adult) | P-value (younger adult vs. frail older adult) | P-value (fit older adult vs. frail older adult) |
| --- | --- | --- | --- | --- | --- | --- |
| Living situation, n (%) |  |  |  | NA | NA | NA |
| Institutionalized | - | 0 (0) | 0 (0) |  |  |  |
| At home | - | 19 (100) | 17 (94.4) |  |  |  |
| Alone | - | 7 (36.8) | 6 (35.4) |  |  |  |
| With others | - | 12 (63.2) | 11 (64.7) |  |  |  |
| Missing | - | 0 (0) | 1 (5.6) |  |  |  |
| Risk of malnutrition, n (%) | - | 10 (52.6) | 11 (61.1) | NA | NA | NA |
| History of delirium, n (%) | - | 0 (0) | 2 (11.8) | NA | NA | NA |
| History of dementia, n (%) | - | 0 (0) | 1 (5.9) | NA | NA | NA |
| Cognitive impairment<sup>b</sup>, n (%) | - | 0 (0) | 6 (35.3) | NA | NA | NA |
| Fall past 6 months, n (%) | - | 0 (0) | 13 (72.2) | NA | NA | NA |
| ADL dependent<sup>c</sup>, n (%) | - | 0 (0) | 2 (11.8) | NA | NA | NA |
| iADL dependent<sup>d</sup>, n (%) | - | 0 (0) | 10 (58.8) | NA | NA | NA |
| **General characteristics** |  |  |  |  |  |  |
| Age (years), mean (SD) | 58.5 (7.7) | 75.7 (5.2) | 76.6 (4.9) | <0.001<sup>‡</sup> | <0.001<sup>‡</sup> | 0.393<sup>‡</sup> |
| Male gender, n (%) | 30 (85.7) | 17 (89.5) | 13 (72.2) | 0.695<sup>‡</sup> | 0.235<sup>‡</sup> | 0.181<sup>‡</sup> |
| Charlson Comorbidity Index, median (IQR) | 0 (0-1) | 2 (0-3) | 2 (1-3) | 0.005<sup>‡</sup> | <0.001<sup>‡</sup> | 0.373<sup>‡</sup> |
| Number of medications, mean (SD) | 3.3 (3.7) | 4.5 (3.2) | 6.3 (4.1) | 0.118<sup>‡</sup> | 0.007<sup>‡</sup> | 0.174<sup>‡</sup> |
| BMI (kg/m<sup>2</sup>), mean (SD) | 26.0 (3.6) | 26.0 (3.9) | 26.6 (4.3) | 0.760<sup>‡</sup> | <0.001<sup>‡</sup> | <0.001<sup>‡</sup> |
| Smoking, n (%) |  |  |  | 0.046<sup>#</sup> | 0.066<sup>#</sup> | 0.909<sup>#</sup> |
| Current or history | 26 (74.3) | 13 (68.4) | 12 (66.7) |  |  |  |
| Alcohol, n (%) |  |  |  | 0.056<sup>‡</sup> | 0.607<sup>‡</sup> | 0.051<sup>‡</sup> |
| Current or history | 27 (77.1) | 16 (88.9) | 10 (55.6) |  |  |  |
| Unknown | 0 (0) | 1 (5.3) | 0 (0) |  |  |  |
| **Disease-specific** |  |  |  |  |  |  |
| New primary tumor, n (%) | 35 (100) | 19 (100) | 18 (100) | NA | NA | NA |
| cT-stage, n (%) |  |  |  | <0.001<sup>‡</sup> | <0.001<sup>‡</sup> | 0.667<sup>‡</sup> |
| Tx | 0 (0) | 2 (10.5) | 1 (5.6) |  |  |  |
| T1 | 0 (0) | 1 (5.3) | 0 (0) |  |  |  |
| T2 | 3 (8.6) | 9 (47.4) | 9 (50) |  |  |  |
| T3 | 32 (91.4) | 5 (26.3) | 8 (44.4) |  |  |  |
| T4 | 0 (0) | 2 (10.5) | 0 (0) |  |  |  |
| cN-stage, n (%) |  |  |  | 0.004<sup>‡</sup> | 0.208<sup>‡</sup> | 0.196<sup>‡</sup> |
| N0 | 8 (22.9) | 11 (57.9) | 7 (38.9) |  |  |  |
| N1 | 12 (34.3) | 6 (31.6) | 7 (38.9) |  |  |  |
| N2 | 15 (42.8) | 2 (10.5) | 2 (11.1) |  |  |  |
| N3 | 0 (0) | 0 (0) | 2 (11.1) |  |  |  |
| M-stage, n (%) |  |  |  | 0.485<sup>‡</sup> | 0.232<sup>‡</sup> | 0.151<sup>‡</sup> |
| Mx/M0 | 34 (97.1) | 19 (100) | 16 (88.9) |  |  |  |
| M1 | 1 (2.9) | 0 (0) | 2 (11.1) |  |  |  |

Abbreviations: SD, Standard deviation; n, number; IQR, Interquartile range; BMI, Body Mass Index; NA, not applicable; ADL, Activities of Daily Living; iADL, Instrumental Activities of Daily Living.
Missing data were not accounted for in the frequencies. Missing values per variable: 6CIT n = 1 (frail n = 1), living situation n = 1 (frail n = 1), history of delirium n = 1 (frail n = 1), history of dementia n = 1 (frail n = 1), ADL n = 1 (frail n = 1), iADL n = 2 (fit n = 1, frail n = 1).
<sup>‡</sup>Mann-Whitney U test
<sup>‡</sup>Fisher's exact test
<sup>#</sup>Chi-squared test

The mean chronological age of the younger adults was 58.5 years, 75.7 years of the fit older adults, and 76.6 years of the frail older adults. The chronological age of the fit and frail older adults was not statistically significantly different (*P* = 0.393). In addition, younger adults had significantly fewer comorbidities (Charlson Comorbidity Index (CCI) of 0) than fit older adults (CCI of 2, *P* = 0.005) and frail older adults (CCI of 2, *P* <0.001). However, the CCI was not significantly different between the fit and frail older adults (*P* = 0.373). Frail older adult patients had a higher BMI than fit older adult patients (mean 26.6 vs. 26.0 kg/m^2^, *P* <0.001). The number of medications used (*P* = 0.007) and BMI (*P* <0.001) were significantly higher in the frail older adult group versus the younger adult group but were not significantly different between the fit older adult group and the younger adult group. Concerning clinical tumor characteristics, the clinical T stage was significantly different between the younger adult and the separate older adult groups (*P* <0.001). However, we observed no significant differences in clinical T (*P* = 0.667) or N (*P* = 0.196) stage between the fit and frail older adult groups. Clinical N stage was significantly different between the younger adult and fit older adult group (*P* = 0.004), but not between the younger adult and frail older adult group (*P* = 0.208).

The pathology slides of pre-treatment esophageal biopsy tissues were assessed on intratumoral stroma quantity, as scored by the histologic tumor-stroma ratio (TSR), and quantification of the tumor immune cell infiltrate (TICI). Illustrative images of stroma-low and stroma-high tumors can be found in *Figure 1A-B*. A total of 8 (11.1%) slides required a third review by an independent observer to reach a complete agreement. We observed stroma-high tumors in 29.2% (*n* = 21) of the patients and stroma-low tumors in 70.8% (*n* = 51) of the patients in our cohort. Subsequently, the TICI within the stromal compartment was quantified. Illustrative images of TICI scoring can be found in *Figure 1C-D*. A total of 13 (18.1%) slides required a third review by an independent observer to reach a complete agreement. The Cohen’s kappa coefficient for interobserver variability was 0.86, indicating near-perfect agreement. Given the current lack of a standardized cut-off value, tumors were categorized into immune-high (>40% stromal infiltration) and immune-low (≤40% stromal infiltration) based on the median TICI percentage (40%) in our cohort. We observed immune-high and immune-low tumors in 43.1% (*n* = 31) and 56.9% (*n* = 41) of the cases, respectively.

**Figure 1.**
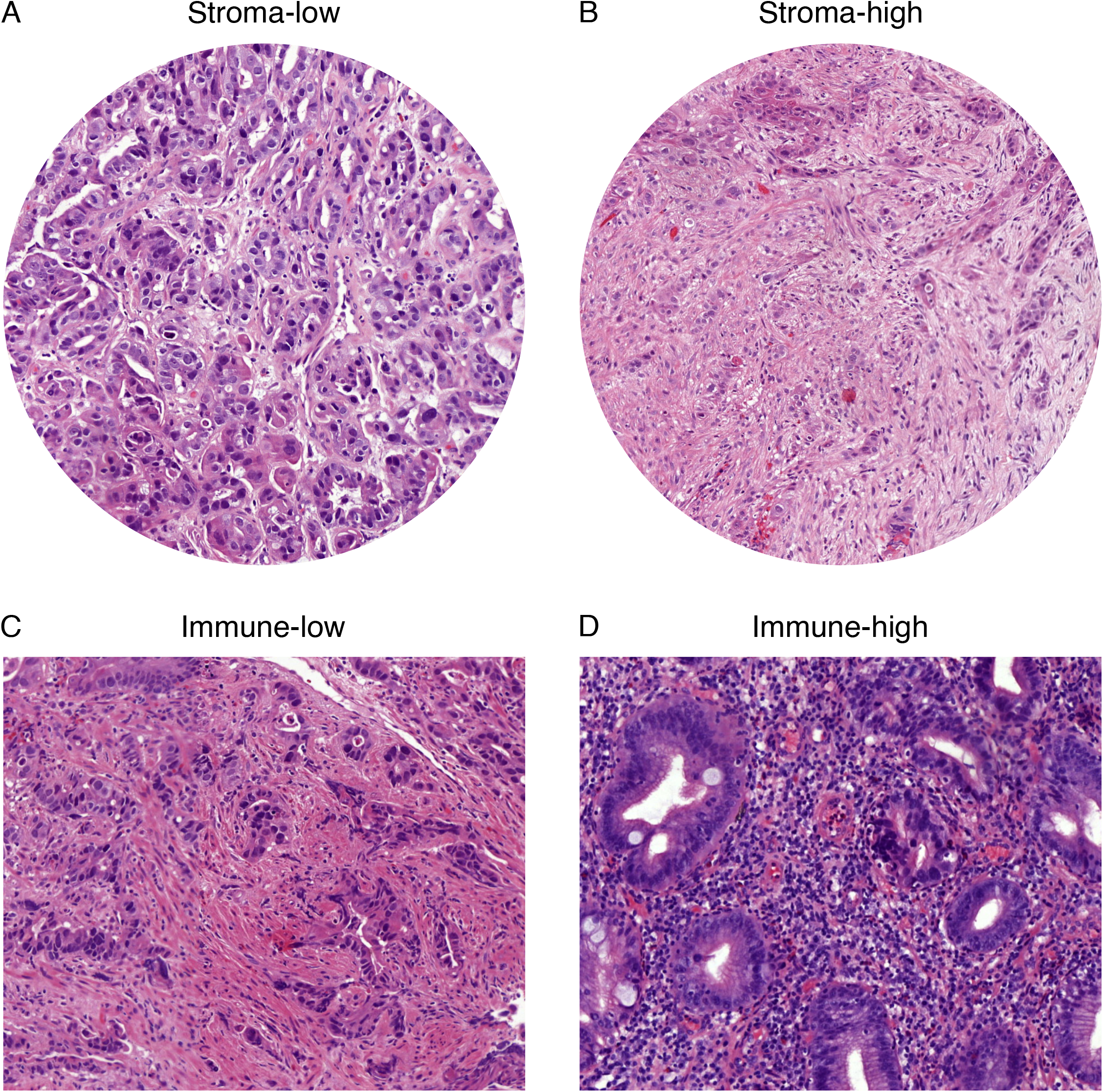
Histopathologic tumor microenvironment features of esophageal adenocarcinoma. Illustrative images of a (A) stroma-low tumor and a (B) stroma-high tumor. Illustrative images of a tumor with (C) low stromal immune cell infiltration and (D) high stromal immune cell infiltration.

### Frailty is Associated with Decreased Intratumoral Stroma Quantity

Next, the association between the intratumoral stroma quantity and the biological age subpopulations was evaluated. We first studied the association between chronological age and intratumoral stroma quantity and found no significant differences in stroma percentage between the younger adult (median 50%, IQR 30-60%) and the total older adult (median 40%, IQR 30-50%) group (*P* = 0.080; *Figure 2A*). Chronological age was not significantly correlated to intratumoral stroma quantity (*Rho* = -0.192, *P* = 0.105; *Supplementary Figure S2A*). We then performed linear regression, adjusted for the covariates sex, chronological age, clinical T and N stage, to test if a progressing biological age group as an independent variable predicted intratumoral stroma quantity. Using the fit older adults as a reference group, we found that the frail older adult group significantly predicted decreased intratumoral stroma percentage (B = -14.66% stroma, *P* = 0.022), whereas the younger adult group did not (B = -4.981% stroma, *P* = 0.562). Regression statistics can be found in *Supplementary Table S1*. Further analysis demonstrated significant differences in stroma percentage between the younger adult (median 50%, IQR 30-60%) and the frail older adult (median 30%, IQR 20-40%) group (*P* = 0.006) and between the fit (median 40%, IQR 30-60) and the frail (median 30%, IQR 20-40%) older adult group (*P* = 0.014; *Figure 2B*). The stroma percentages of the fit older group (median 40%, IQR 30-60) resembled those of the younger adult group (median 50%, IQR 30-60%; *P* = 0.876). Applying the standardized cut-off value for the TSR (50%), the proportion of stroma-high (>50% stroma) and stroma-low (≤50% stroma) tumors was significantly different between the strata of biological age (*X*^2^ = 8.107, *P* = 0.017; *Supplementary Figure S2B*). Tumors in the younger adult group more often demonstrated a histologic stroma-high phenotype (42.9%) in comparison to tumors in the fit older (26.3%) and frail older adult (5.6%) group.

**Figure 2.**
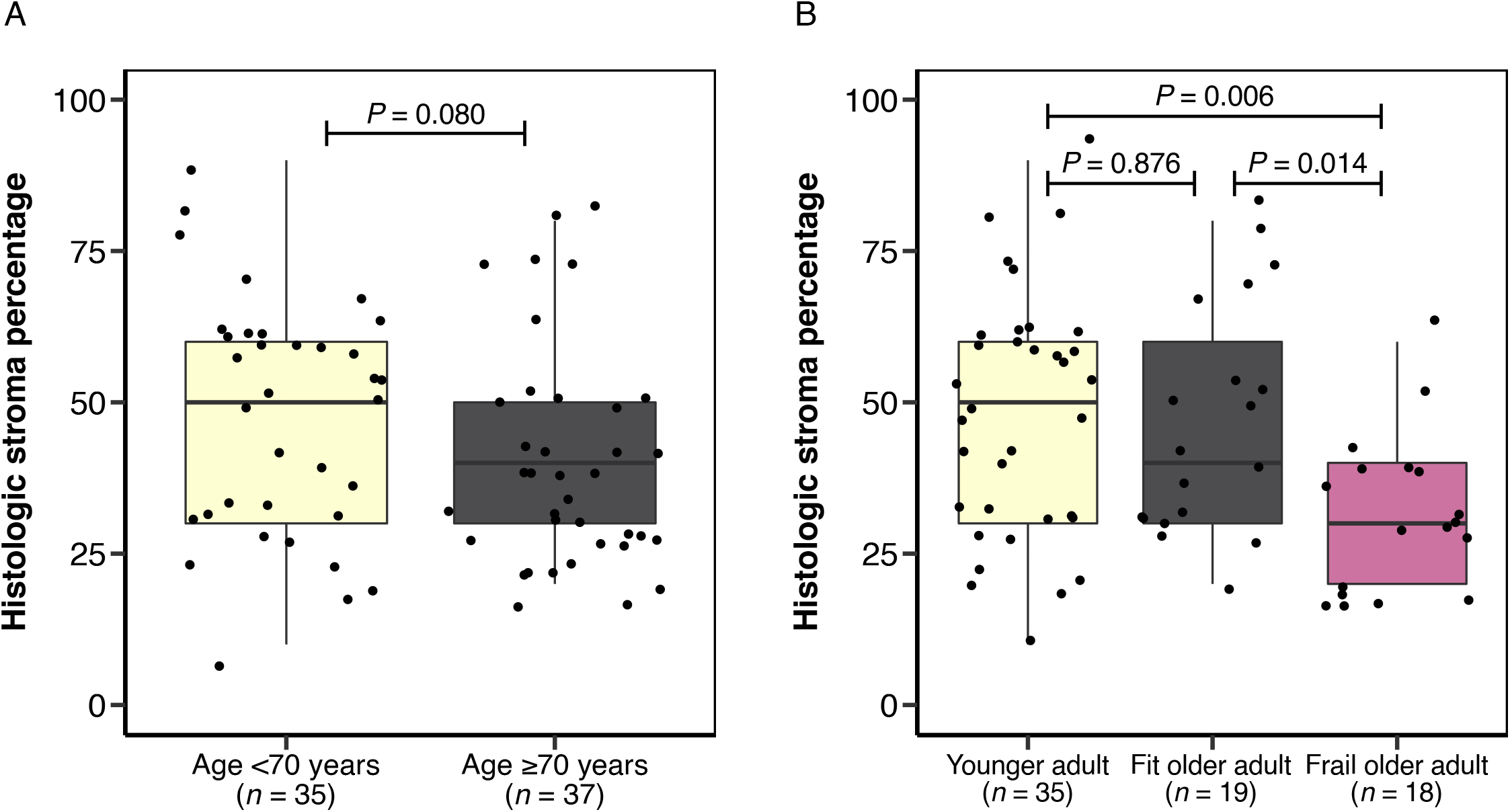
Histologic intratumoral stroma content and age. (A) Boxplot of the histologic intratumoral stroma percentage, as scored by the tumor-stroma ratio, and chronological age. (B) Boxplot of the intratumoral stroma percentage and strata of biological age, as categorized by a geriatric assessment.

### Vast Heterogeneity of the Histologic Tumor Immune Cell Infiltrate in Strata of Biological Age

Next, we studied the association between visually-scored histologic TICI and biological age. The histologic TICI demonstrated vast heterogeneity in the younger adult (median 40%, IQR 30-60%), fit older adult (median 40%, IQR 30-75%), and frail older adult (median 35%, IQR 30-67.5%) groups (*Figure 3*). Consequently, we observed no statistically significant differences in TICI between the younger adult and fit older adult group (*P* = 0.301), the younger adult and frail older adult group (*P* = 0.389), and the fit and frail older adult group (*P* = 0.969; *Figure 3*). In addition, histologic TICI was not correlated to chronological age (*Rho* = 0.070, *P* = 0.558; *Supplementary Figure S3*).

**Figure 3.**
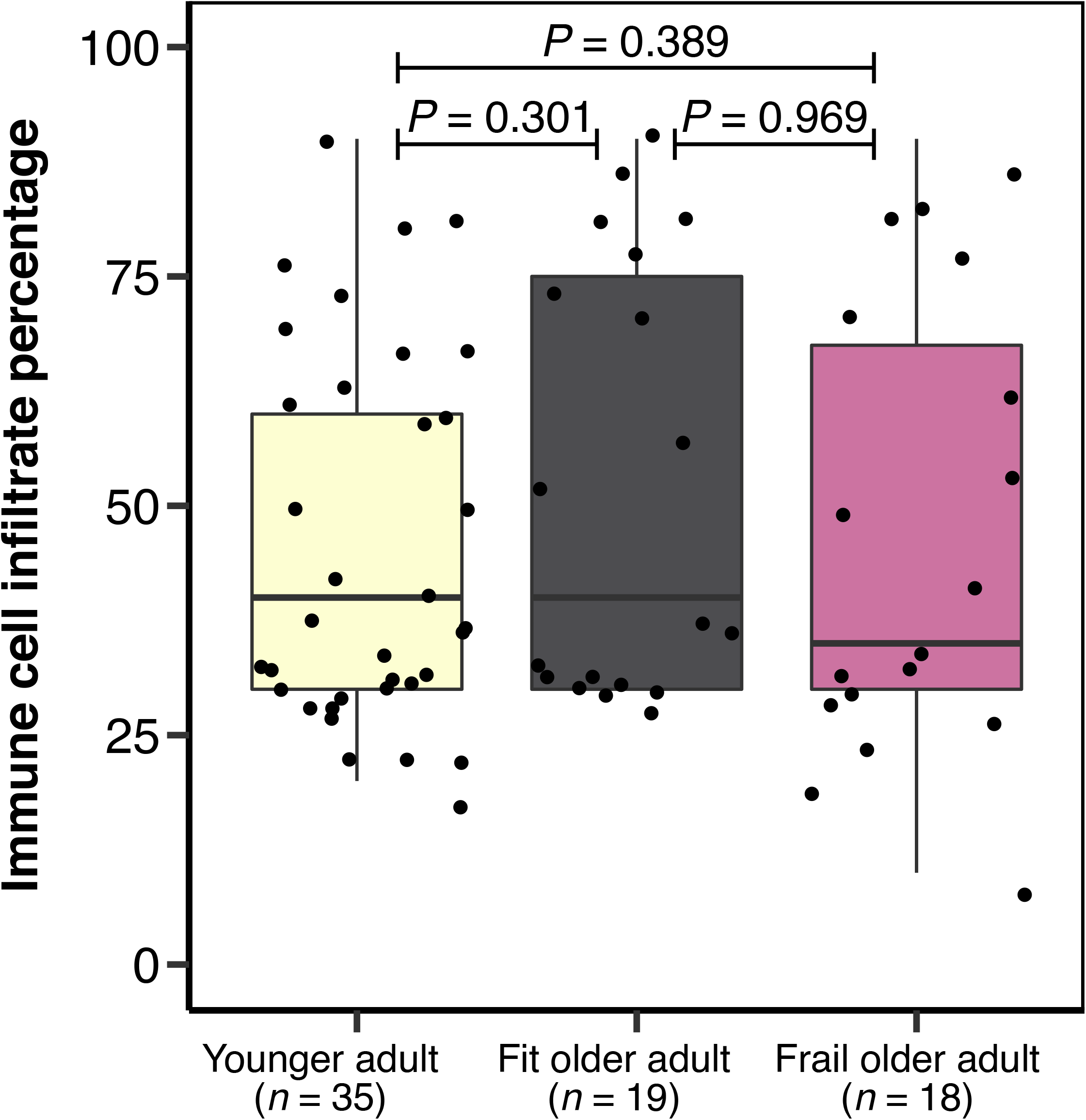
Boxplot of histologic tumor immune cell infiltrate (TICI) and strata of biological age.

### Tumors in Frail Older Adults Are Associated with a Desert Tumor Microenvironment Phenotype

We then combined quantification of the histologic intratumoral stroma quantity and the TICI into a single categorical parameter for histologic TME phenotype. Tumors were subsequently categorized as stroma-low/immune-low (SLIL), stroma-low/immune-high (SLIH), stroma-high/immune-low (SHIL), and stroma-high/immune-high (SHIH). Interestingly, the proportion of tumors with a desert TME phenotype (SLIL) successively increased in the younger adult (34.3%), fit older adult (36.8%), and frail older adult (55.6%) group (*Figure 4*). In contrast, the proportion of SHIL tumors, a TME phenotype associated with poor treatment response, markedly decreased in the successive younger adults (25.7%), fit older (15.8%), and frail older (0%) groups (35, 36). Despite these trends, the histologic TME phenotype distribution was statistically significantly different between the younger adult and frail older adult group (*X*^2^ = 8.212, *P* = 0.042), but not between the fit older adult and frail older adult group (*X*^2^ = 3.84, *P* = 0.279), likely due to small group numbers. In addition, we observed no statistically significant difference in histologic TME phenotype between the younger adult and fit older adult group (*X*^2^ = 1.800, *P* = 0.615).

**Figure 4.**
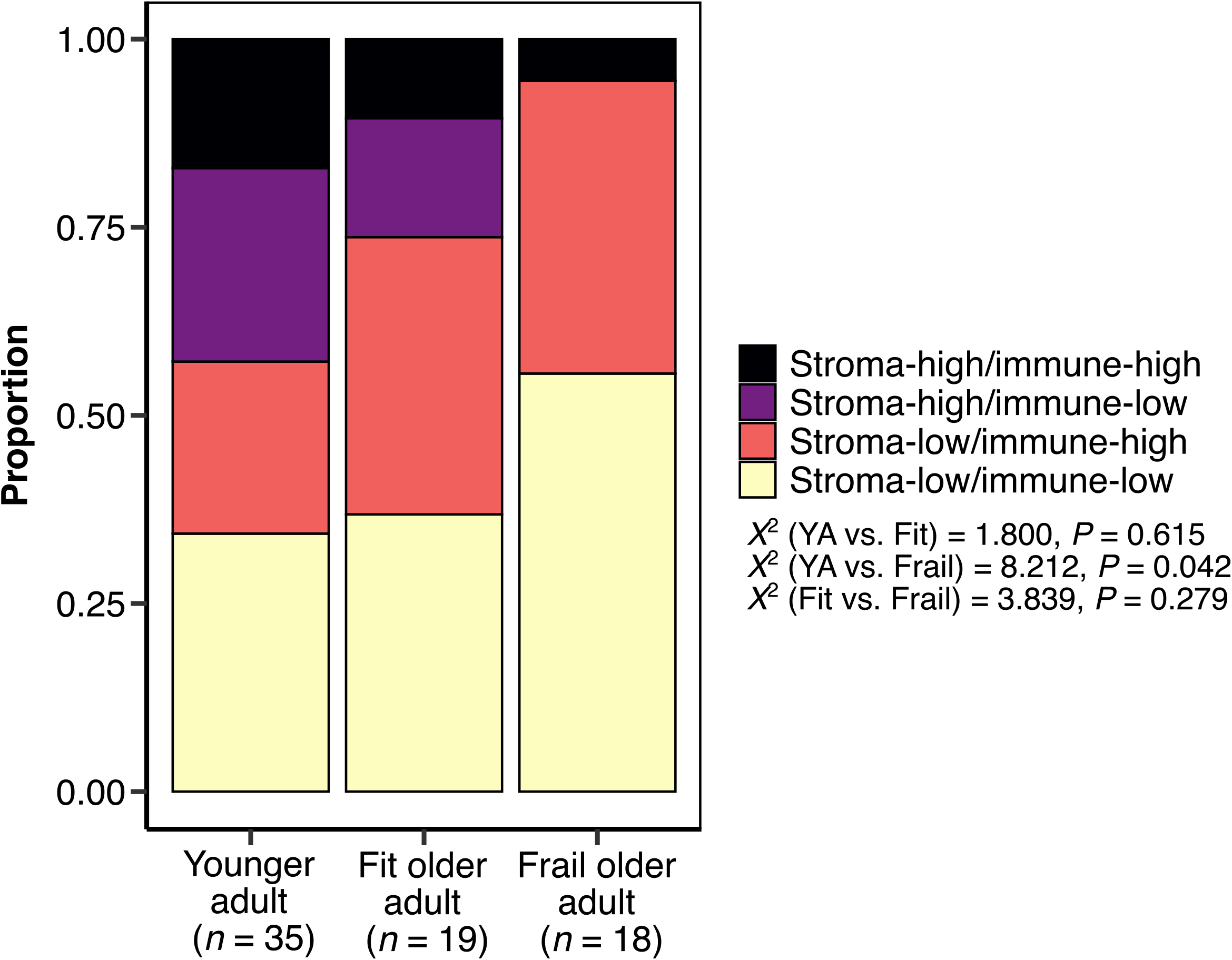
Proportion of histologic tumor microenvironment phenotypes, composed of histologic intratumoral stroma percentage and tumor immune cell infiltrate, in strata of biological age, as categorized by a geriatric assessment. SHIH, stroma-high/immune-high; SHIL, stroma-high/immune-low; SLIH, stroma-low/immune-high; SLIL, stroma-low/immune-low; X2, Pearson Chi-square.

### Decreased Blood Thrombocyte Counts in Frail Older Adult Patients

Since we and others recently demonstrated increased transcription of genes associated with thrombocyte activation and increased blood thrombocyte counts in patients with stroma-high tumors, we wondered whether the lower stroma percentages observed in the frail older group were associated with decreased blood thrombocyte counts at the time of diagnosis (20, 37).

Similar to intratumoral stroma quantity, we performed linear regression, adjusted for the covariates sex, chronological age, and clinical T and N stage, to test if a progressing biological age group significantly predicted blood thrombocyte counts. Using the fit older adults as a reference population, both the frail older adult group (B = -44.94 thrombocyte count (10^9^), *P* = 0.089) and the younger adult group (B = 15.77 thrombocyte count (10^9^), *P* = 0.654) did not significantly predict blood thrombocyte count, relative to the fit older adults. Regression statistics can be found in *Supplementary Table S2*. Although no statistically significant results were observed in linear regression, the frail older adult group demonstrated a lower median blood thrombocyte count in comparison to the younger adult (median 295 (10^9^), IQR 256.5-316.5) group (*P* = 0.032; *Figure 5A*). No statistically significant differences were observed in blood thrombocyte counts between the fit older adult (median 255 (10^9^), IQR 204.5-322.5) and frail older adult (median 239 (10^9^), IQR 209-285) group (*P* = 0.486). In addition, we observed no significant correlation between blood thrombocyte count at the time of diagnosis and histologic intratumoral stroma quantity in this study (*Rho* = 0.215, *P* = 0.072; *Supplementary Figure S4*). Blood thrombocyte count was not correlated to chronological age (*Rho* = -0.184, *P* = 0.124).

**Figure 5.**
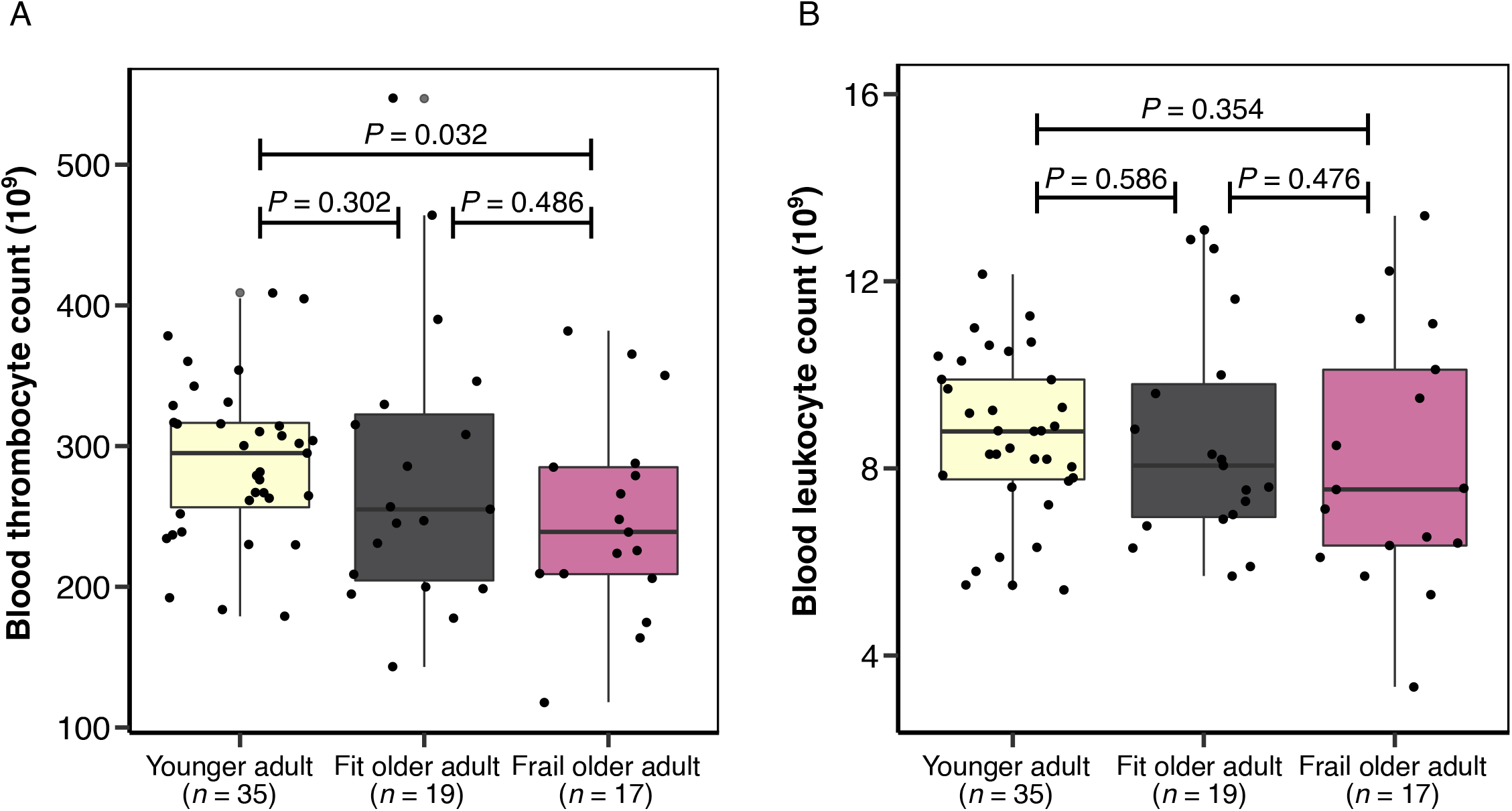
Blood cell count markers and biological age. (A) Boxplot of blood thrombocyte count at the time of diagnosis and strata of biological age. (B) Boxplot of blood leukocyte count at the time of diagnosis and strata of biological age.

In linear regression, using the fit older adults as a reference population, we found that both the frail older adult group (B = -0.838 leukocyte count (10^9^), *P* = 0.276) and the younger adult group (B = -0.865 leukocyte count (10^9^), *P* = 0.402) did not significantly predict blood leukocyte count. Regression statistics can be found in *Supplementary Table S3*. Blood leukocyte count was not statistically significantly different between the younger adult (median 9 (10^9^), IQR 7.8-9.9) and fit older adult (median 8.1 (10^9^), IQR 7.0-9.8) group (*P* = 0.586), the younger adult and frail older adult (median 7.6 (10^9^), IQR 6.4-10.1) group (*P* = 0.354), and the fit and frail older adult group (*P* = 0.476; *Figure 5B*). In addition, no significant correlation was found between blood leukocyte count and histologic TICI (*Rho* = -0.146, *P* = 0.224) and between blood leukocyte count and chronological age (*Rho* = -0.132, *P* = 0.272; *Supplementary Figure S4*).

## Discussion

In the current work, we addressed the association between geriatric assessment (GA) and characteristics of the tumor microenvironment (TME) in patients with esophageal adenocarcinoma with an indication for intensive treatment. Tumors in frail older adults demonstrated lower intratumoral stroma quantity in comparison to tumors in chronological-age-matched fit older adults, independent of tumor stage. In addition, the low intratumoral stroma quantity in frail older adults was accompanied by a decreasing trend in blood thrombocyte count. Lastly, we found an increased proportion of tumors with a histologic desert TME phenotype, comprising low stroma quantity and low immune cell infiltration, in frail older adults.

Biological age aims to account for interindividual heterogeneity in health and can markedly contrast chronological age. In the past recent decades, extensive research efforts have been made to formulate reliable predictors of biological age (8, 38). To this aim, advances in molecular biology have identified associations between patient fitness or frailty and omics data, such as DNA methylation, gene expression profiles, and protein glycosylation (39-42). Despite these findings, there is no consensus biomarker for biological age at this time. As the rate of biological aging varies across different tissues within the same individual, a universally applicable biomarker to capture the overall biological age might not be feasible (43-45). A robust predictor of biological age in clinical practice may therefore be a context-dependent biomarker that can be used in framed clinical settings only. At this time, there is no specific biomarker for assessing biological age in the clinical oncology setting.

The distinct differences in TME intrinsic properties between biological age groups observed here may ultimately serve as a biomarker for the identification of oncologic frail older adult patients. To this date, only few studies have elaborated on the relation between the TME and biological age, whereas multiple studies considering the chronologically aging TME report conflicting results (15, 46-51). In a recent study by our group, we observed increased intratumoral stroma quantity in progressive chronological age groups (70-<75, 75-<80, 80-<85, 85-<90, and ≥90 years) in patients with breast cancer (46). The observation of increased intratumoral stroma quantity in older adult patients with breast cancer was previously validated by an independent research group (47). In contrast, multiple reports on breast cancer patients failed to demonstrate an association between intratumoral stroma quantity and chronological age (48, 49). Likewise, intratumoral stroma quantity was not associated with chronological age in esophageal cancer (15, 50, 51). In the current work, we did not observe an association between intratumoral stroma quantity and chronological age but demonstrated significant differences when chronological age groups were further stratified by a clinical parameter for biological age. In light of these findings, the prior analyses of intratumoral stroma quantity and chronological age may therefore potentially be confounded by skewed biological age distributions within the respective study cohorts.

In addition to distinct histologic TME features, we observed a decline in blood thrombocyte counts in the successive biological age groups included in this study. Blood thrombocyte counts were previously shown to decrease with advancing chronological age (52, 53). Interestingly, the declining blood thrombocyte counts demonstrated a similar trend to the decrease in intratumoral stroma quantity across the biological age groups. Tumor-infiltrating thrombocytes and coagulation factors are key constituents of the TME and impact tumor progression and drug resistance (54-56). We recently reported increased transcription of genes associated with thrombocyte degranulation in stroma-high colon tumors (37). In addition, a recent report on breast cancer described increased thrombocyte-associated protein expression in stroma-high tumors. In subsequent blood analyses, the authors observed increased blood thrombocyte counts in patients with stroma-high tumors (20). Likewise, we observed decreased intratumoral stroma quantity and blood thrombocyte counts in frail older adult patients. Although we did not observe a statistically significant correlation between blood thrombocyte count and stroma quantity in this study, it is plausible that blood thrombocyte count may reflect histologic stroma quantity and, hence, be informative as a blood biomarker for TME characterization.

We found decreased histologic intratumoral stroma quantity, a phenotype associated with low stromal cell abundance, in tumors of frail older adults relative to tumors of fit older adults. This observation may suggest an increased proliferative arrest of TME stromal cell populations in tumors of frail older adults. Paradoxically, while age-related changes induce growth arrest (cellular senescence), cell loss (apoptosis), and functional degradation of stromal and immune cell populations, senescence-associated changes in TME cell populations were found to promote tumor progression and metastasis (24). This conflicting phenomenon is captured by the term antagonistic pleiotropy, a trait that is beneficial in early life may be detrimental at an older age (57). One of the key features thought to be responsible for this phenomenon is the acquisition of a senescence-associated secretory phenotype (SASP) of TME cell populations, a distinct secretory profile of senescent cells that demonstrates both pro-tumorigenic and tumor-suppressive properties (58). The SASP is characterized by the secretion of extracellular matrix (ECM) degrading factors, such as matrix metalloproteinases, suggesting ECM catabolism, and age-related stromal cell senescence has been shown to induce local anti-fibrotic effects (59-61). Interestingly, the authors of a study on breast cancer reported an increased expression of senescence-associated genes in the intratumoral stroma of patients aged ≥80 years old in comparison to a young reference population aged <45 years old (62). In addition, a recent study on aging and the SASP secretome observed a positive association between circulating SASP protein abundance and clinical patient frailty (63). In conjunction with these reports, we observed a large fraction of tumors with a desert histologic TME phenotype (i.e., low stroma quantity and low immune cell infiltration) in the frail older adult patient population, illustrative of the biological age-related degenerative adaptations. Our observation may therefore imply an increased SASP of the local TME cell populations in comparison to the populations in tumors of fit older adults. Although the stable cell cycle arrest of senescent cells is presumed to contribute to tumor suppression in later life, the chronic inflammatory characteristics of the SASP secretome have paradoxically been shown to promote tumorigenesis (64). At this time, it remains unclear how our biological age-associated histologic findings relate to clinical tumor aggressiveness, which should be the topic of future studies.

Of note, the contrasts in intratumoral stroma quantity and blood thrombocyte counts between younger adult and older adult patients were more pronounced when the older population was further categorized into fit and frail older adult subpopulations. Comprehensive geriatric assessment is widely accepted as the golden standard for determining older adult patient frailty (65). Our findings provide a biological underpinning for the clinical relevance of assessing frailty, further justifying the need for standardized geriatric assessment in geriatric cancer patients. Recent randomized-controlled trials have shown that comprehensive geriatric assessment may result in chemotherapy dose reduction and overall lower chemo-toxicity in geriatric patients (66, 67). Patient selection by geriatric assessment may therefore reflect clinically-relevant biological differences between fit and frail older adults, amongst which the tumor microenvironment differences described here. Although we were not able to assess tumor prognosis and aggressiveness in our study, our observation of decreased intratumoral stroma quantity and decreased blood thrombocyte counts in frail older adults may suggest a decreased potential of metastasis and aggressiveness of esophageal tumors in this patient population. Interestingly, a large proportion of tumors in the frail older population exhibited low stroma quantity and high immune cell infiltrates, a TME phenotype that is associated with favorable (immuno)therapy response (35, 36). This latter observation may suggest overall increased responsivity to cancer treatment regimens in the frail older adult patient population, supporting the notion that frail older adult patients with esophageal cancer are likely to benefit from biological age-tailored treatment regimens entailing less intensive agents (12, 25). Importantly though, in this study, we were not able to discriminate pre-existent frailty from oncology-induced frailty. Since the latter is believed to be the result of increased disease activity, oncology-induced frailty would imply a more aggressive tumor phenotype and thus poorer survival and treatment outcomes (68-70). Hence, our results warrant a larger cohort follow-up study to validate the histologic TME features in frail older adults and whether these features are indeed associated with poor patient survival and treatment response.

Strengths of the current study include the use of a standardized GA, including the four geriatric domains, to address patient frailty. In addition, the current work comprises the first study, to the best of our knowledge, to associate histologic TME characteristics with clinical patient frailty in older adults with esophageal cancer. Nevertheless, we want to acknowledge the limitations of our study. First, we performed this pilot study on a small sample size of patients that were included in the TENT study. Subsequently, we were not able to create patient groups with equal clinical baseline characteristics. We, therefore, refrained from prognostic analyses, which currently limits the causal interpretability of our TME findings in frail older adults. Although we do believe our results are valid, our observations may still have been confounded by unaccounted differences in baseline characteristics. Therefore, validation of our results in a larger study that accounts for baseline differences is warranted.

The current work demonstrates biological age-related differences in histologic TME phenotypes in esophageal adenocarcinoma. Frailty in older adult patients was associated with decreased histologic intratumoral stroma quantity in comparison to chronological-age-matched fit older adults. In addition, a large proportion of the tumors in frail older adult patients demonstrated a desert histologic TME phenotype, consisting of low stroma quantity and low immune cell infiltration. These frailty-associated histologic findings were accompanied by a coinciding observation of decreased blood thrombocyte counts in frail older adults, a feature that was previously associated with decreased TME activity (20, 54). Despite the presence of methodological limitations in our study, our results illustrate the stromal-reprogramming effects of biological age and subsequently demonstrate how GA may select for older patient subpopulations with distinct TME features. Given that the TME significantly influences therapeutic response and clinical patient outcome in cancer, the application of GA in older adult patients and, hence, the selection of TME subtype, may guide treatment decisions in the older adult patient population in clinical practice.

## Data Availability

All data produced in the present study are available upon reasonable request to the authors.

## Abbreviations

6CIT: 6-item cognitive impairment test
ADL: activities of daily living
BMI: body mass index
CCI: Charlson Comorbidity Index
ECM: extracellular matrix
GA: geriatric assessment
H&E: hematoxylin and eosin
iADL: instrumental activities of daily living
IQR: interquartile range
SASP: senescence-associated secretory phenotype
SD: standard deviation
SHIH: stroma-high/immune-high
SHIL: stroma-high/immune-low
SLIH: stroma-low/immune-high
SLIL: stroma-low/immune-low
TICI: tumor immune cell infiltrate
TME: tumor microenvironment
TSR: tumor-stroma ratio.

## Statements and Declarations

## Acknowledgments

We are greatly indebted to the patients participating in the study. This work was supported by grants from the Bollenstreekfonds, Lisse-Hillegom; the Institute for Evidence-Based Medicine in Old Age (IEMO), and the Vitality Oriented Innovations for the Lifecourse of the Ageing Society (VOILA) project. VOILA is funded by ZonMw (project number 457001001). C.J.R is funded by an MD/PhD grant from the Leiden University Medical Center (LUMC). None of the above parties has had a role in the design of the study, collection, analysis and interpretation of data, and writing of the manuscript.

## Data availability statement

Inquiries concerning data requests can be directed to the corresponding author. The data of this study are available upon reasonable request.

