## Supplementary Figures * Tables for "Association of Biological Age with Tumor Microenvironment in Patients with Esophageal Adenocarcinoma"

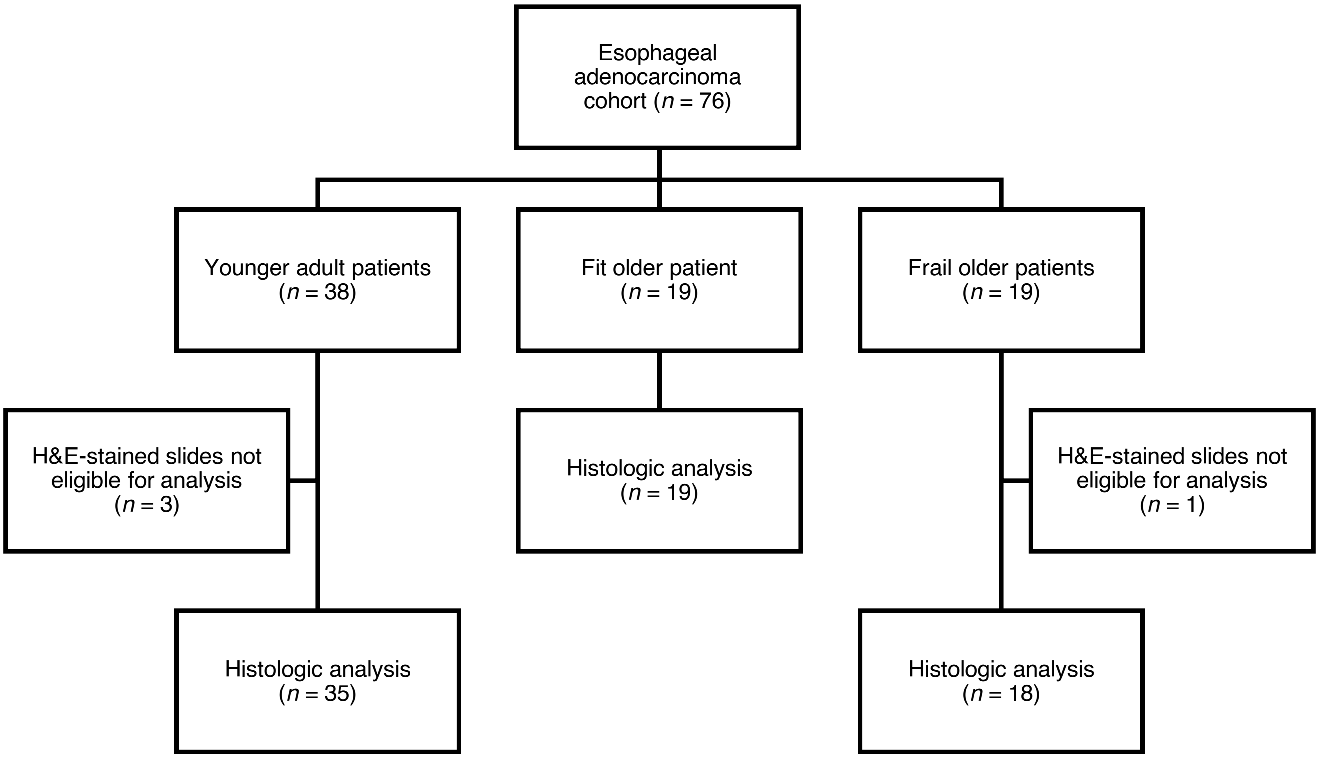


**Supplementary Figure S1.** Flowchart of cohort selection. Out of the 76 patients with esophageal adenocarcinoma selected for this study, 72 were eligible for histologic analysis and subsequently included.

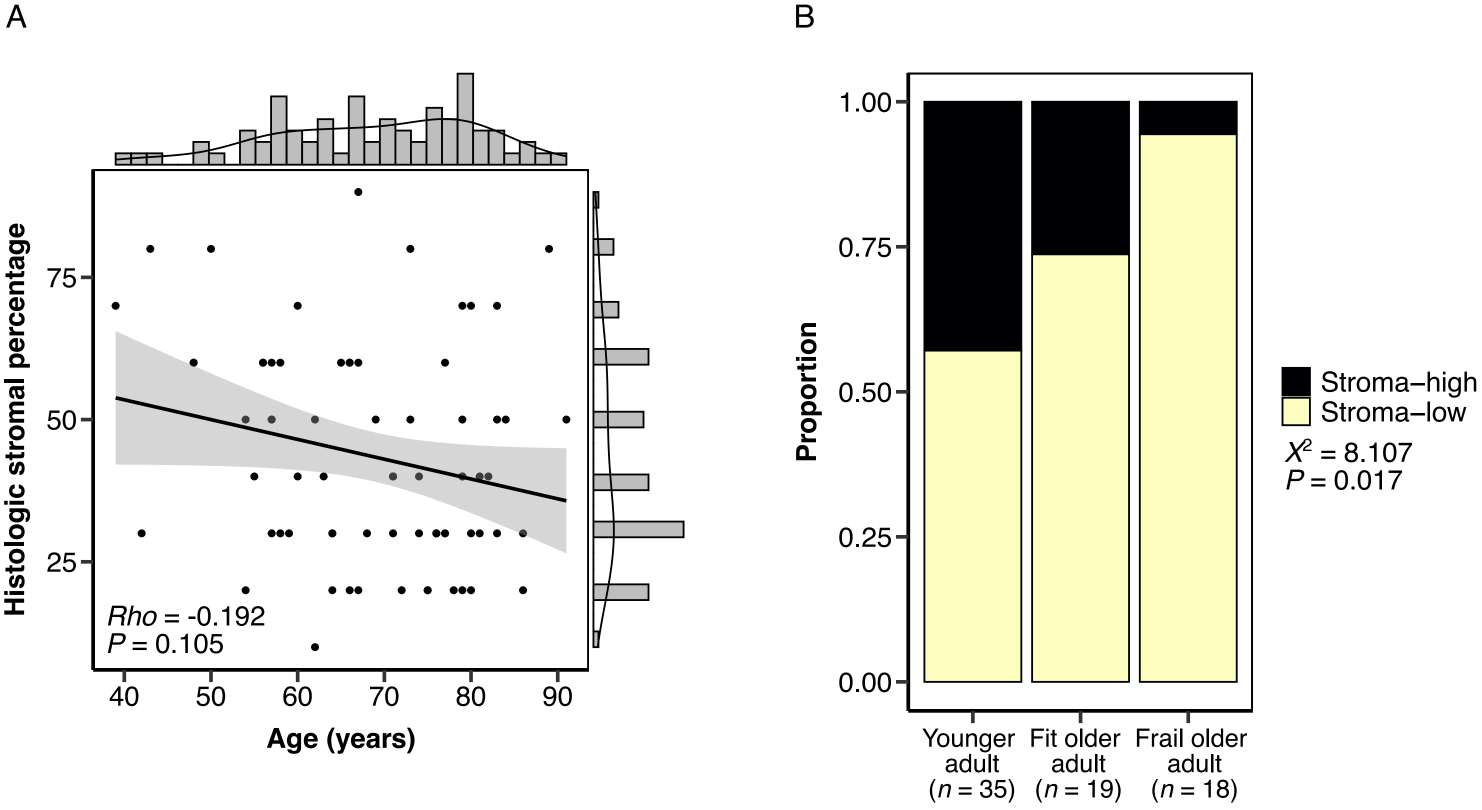


**Supplementary Figure S2.** Histologic intratumoral stroma percentage and age. (A) Scatter correlation plot of histologic intratumoral stroma percentage and chronological age. (B) Stacked bar plot of the proportions of stroma-high and stroma-low tumors, as scored by the standardized histologic tumor-stroma ratio, in strata of biological age.

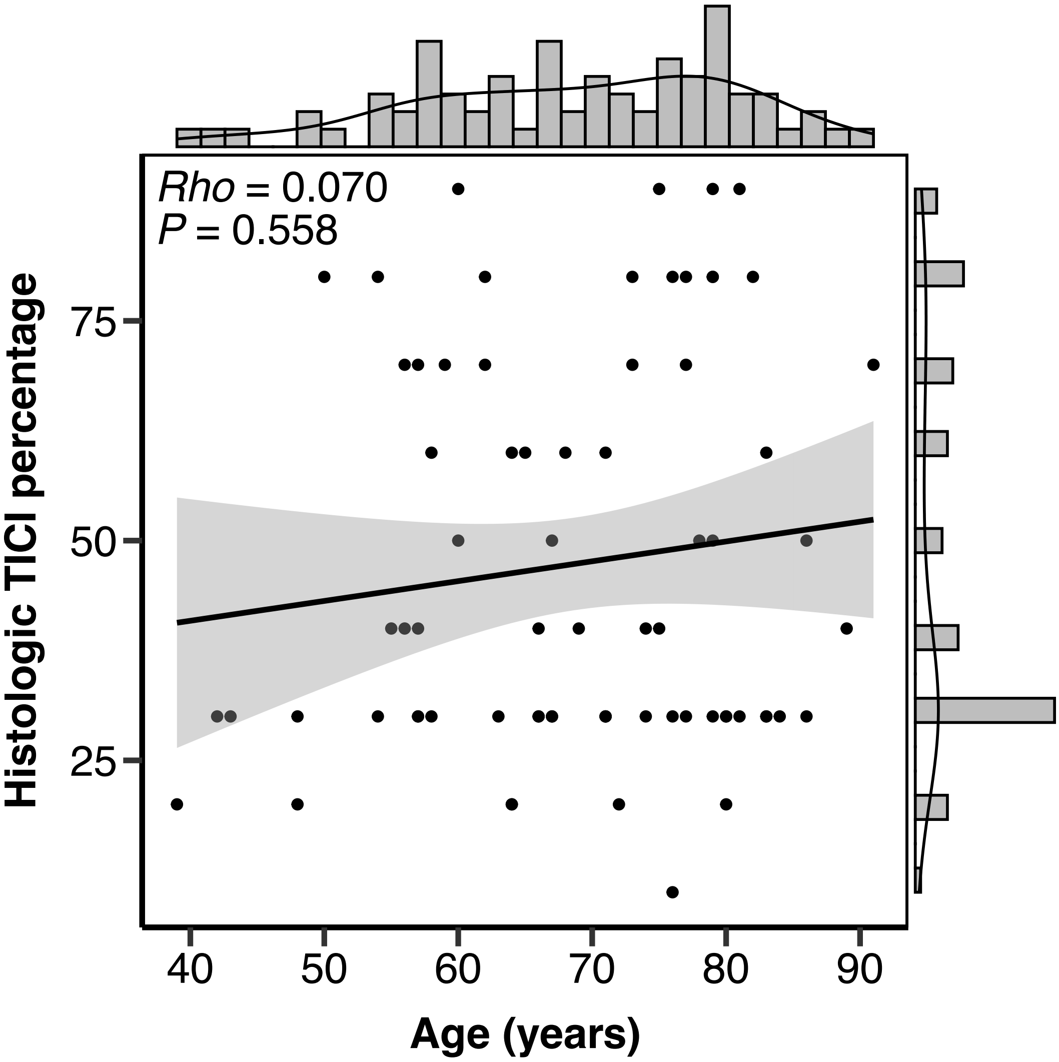


**Supplementary Figure S3.** Scatter correlation plot of histologic tumor immune cell infiltrate (TICI) and chronological age.

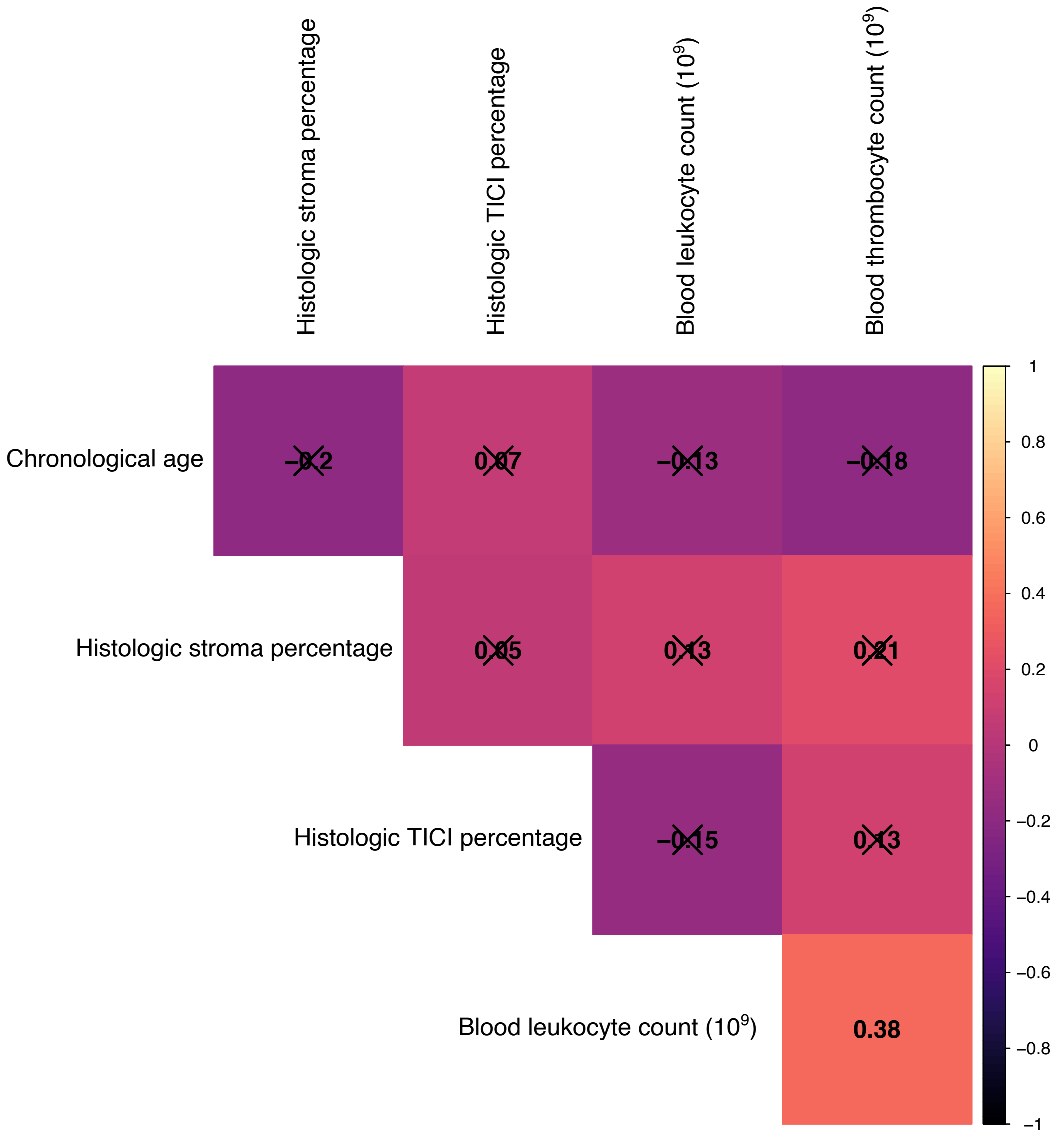


**Supplementary Figure S4.** Spearman correlation coefficient matrix of chronological age, histologic parameters, and blood cell counts. *X, not significant.*

**Supplementary Table S1.** Statistics for a multiple linear regression model containing strata of biological age, chronological age, male sex, clinical T and N stage as predictor variables and histologic intratumoral stroma percentage as dependent variable. The fit older adult group was selected as a reference.

| **Variables** | **Coefficient (B)** | **SE** | **95% CI** | **T statistic** | ***P*-value** |
|---|---|---|---|---|---|
| Constant | 64.02 | 31.30 | 1.500 to 126.5 | 2.045 | 0.045* |
| Younger adult | -4.981 | 8.551 | -22.06 to 12.10 | -0.583 | 0.562 |
| Frail older adult | -14.66 | 6.233 | -27.11 to -2.213 | -2.352 | 0.022* |
| Chronological age | -0.251 | 0.356 | -0.961 to 0.460 | -0.704 | 0.484 |
| Male sex | 0.473 | 5.960 | -11.43 to 12.38 | 0.079 | 0.937 |
| cT stage | 0.998 | 3.295 | -5.583 to 7.579 | 0.303 | 0.763 |
| cN stage | -0.215 | 2.803 | -5.813 to 5.383 | -0.077 | 0.939 |

$R^2_{adj}$ = 0.051, *P* = 0.151
Abbreviations: SE, standard error; CI, confidence interval.
*Statistically significant at *P*<0.05, two-tailed

**Supplementary Table S2.** Statistics for a multiple linear regression model containing strata of biological age, chronological age, male sex, clinical T and N stage as predictor variables and blood thrombocyte count at the time of diagnosis as dependent variable. The fit older adult group was selected as a reference.

| **Variables** | **Coefficient (B)** | **SE** | **95% CI** | **T statistic** | ***P*-value** |
|---|---|---|---|---|---|
| Constant | 248.0 | 128.0 | -7.732 to 503.6 | 1.937 | 0.057 |
| Younger adult | 15.77 | 35.01 | -54.16 to 85.71 | 0.451 | 0.654 |
| Frail older adult | -44.94 | 26.06 | -97.00 to 7.114 | -1.725 | 0.089 |
| Chronological age | 0.821 | 1.455 | -2.085 to 3.728 | 0.565 | 0.574 |
| Male sex | -32.76 | 24.50 | -81.71 to 16.18 | -1.337 | 0.186 |
| cT stage | -4.651 | 13.47 | -31.55 to 22.25 | -0.345 | 0.731 |
| cN stage | 15.99 | 11.56 | -7.110 to 39.10 | 1.383 | 0.172 |

$R^2_{adj}$ = 0.010, *P* = 0.363
Abbreviations: SE, standard error; CI, confidence interval.
*Statistically significant at *P*<0.05, two-tailed

**Supplementary Table S3.** Statistics for a multiple linear regression model containing strata of biological age, chronological age, male sex, clinical T and N stage as predictor variables and blood leukocyte count at the time of diagnosis as dependent variable. The fit older adult group was selected as a reference.

| **Variables** | **Coefficient (B)** | **SE** | **95% CI** | **T statistic** | ***P*-value** |
|---|---|---|---|---|---|
| Constant | 12.08 | 3.746 | 4.599 to 19.57 | 3.225 | 0.002* |
| Younger adult | -0.865 | 1.025 | -2.912 to 1.182 | -0.884 | 0.402 |
| Frail older adult | -0.838 | 0.763 | -2.361 to 0.686 | -1.098 | 0.276 |
| Chronological age | -0.034 | 0.043 | -0.119 to 0.051 | -0.797 | 0.428 |
| Male sex | -0.376 | 0.717 | -1.809 to 1.057 | -0.524 | 0.602 |
| cT stage | -0.325 | 0.394 | -1.113 to 0.462 | -0.825 | 0.412 |
| cN stage | 0.549 | 0.339 | -0.127 to 1.226 | 1.623 | 0.110 |

$R^2_{adj}$ = -0.017, *P* = 0.571
Abbreviations: SE, standard error; CI, confidence interval.
*Statistically significant at *P*<0.05, two-tailed
